# Prediction of antidepressant side effects in the Genetic Link to Anxiety and Depression Study

**DOI:** 10.1101/2024.05.01.24306668

**Authors:** Danyang Li, Yuhao Lin, Helena L. Davies, Johan Källberg Zvrskovec, Rujia Wang, Chérie Armour, Ian R. Jones, Andrew M. McIntosh, Nathalie Kingston, John R. Bradley, Christopher Hübel, Gursharan Kalsi, Jonathan R. I. Coleman, the NIHR BioResource consortium, Matthew Hotopf, Thalia C. Eley, Evangelos Vassos, Raquel Iniesta, Gerome Breen

## Abstract

Antidepressants are the most common treatment for moderate or severe depression. Side effects are crucial indicators for antidepressants, but their occurrence varies widely among individuals. In this study, we leveraged genetic and medical data from self-reported questionnaires in the Genetic Links to Anxiety and Depression (GLAD) study to build prediction models of side effects and subsequent discontinuation across three antidepressant classes (SSRI, SNRI, tricyclic antidepressant (TCA)) at the first and the last (most recent) year of prescription. We included 259 predictors spanning genetic, clinical, illness, demographic, and antidepressant information. Six prediction models were trained, and their performance was compared. The final dataset comprised 4,354 individuals taking SSRI in the first prescription and 3,414 taking SSRI, SNRI or TCA in the last year of prescription. In the first year, the best area under the receiver operating characteristic curve (AUROC) for predicting SSRI discontinuation and side effects were 0.65 and 0.60. In the last year of SSRI prescription, the highest AUROC reached 0.73 for discontinuation and 0.87 for side effects. Models for predicting discontinuation and side effects of SNRI and TCA showed comparable performance. The history of side effects and discontinuation of antidepressant use were the most influential predictors of the outcomes in the last year of prescription. When examining 30 common antidepressant side effect symptoms, most of them were differentially prevalent between antidepressant classes. Our findings suggested the feasibility of predicting antidepressant side effects using a self-reported questionnaire, particularly for the last prescription. These results could contribute valuable insights for the development of clinical decisions aimed at optimising treatment selection with enhanced tolerability but require replication in medical record linkage or prospective data.

## Introduction

Antidepressants are commonly prescribed medications, but both clinicians and their patients face uncertainty and concerns due to a lack of ability to accurately predict who will experience side effects. The majority of individuals taking antidepressants report experiencing at least one side effect^1^. In clinical trials, more than half of individuals discontinue medication within six months of starting treatment, with side effects being the most commonly cited contributing factor^2^. Side effects often result in staged and/or inadequate dosing, which is associated with a lack of remission or delayed remission^1^. Some side effects, such as fatigue, sleep change, and cognitive impairment phenomenologically overlap with depression symptoms and related comorbidities, such as anxiety. This overlap further complicates treatment decisions such as dose adjustment or the selection of appropriate initial or alternative medications.

Tricyclic antidepressants (TCAs) were the dominant medications used to treat depression before the 1990s. Currently, second-generation antidepressants such as selective serotonin reuptake inhibitors (SSRIs) and serotonin-norepinephrine reuptake inhibitors (SNRIs) are first-line treatments for depression. All antidepressants operate by targeting monoamine neurotransmitters to increase activation of postsynaptic receptors^3^. Individuals taking TCAs are more likely to discontinue treatment, to experience lower tolerability and increased cardiac side effects compared to those taking second-generation antidepressants^4,5^. Conversely, side effects such as sexual dysfunction, insomnia, nausea, irritability, and anxiety are more prominent response to SSRIs and SNRIs^6^. Nevertheless, clarity over which side effect symptoms are specific to different antidepressants is lacking.

Within the same type of drugs, there is considerable inter-individual variance in side effect occurrence and severity. Various factors, including sex^7^ and clinical features like depression severity^8^, have been associated with antidepressant tolerability and adherence. Pharmacogenomic testing is an emerging strategy to inform medication selection and dose decisions based on individual genetic variations. Variants such as those found in the cytochrome P450 genes *CYP2C19* and *CYP2D6* are linked to antidepressant metabolism and clearance. They can cause differences in enzyme activity that influence antidepressant response and side effects, although their reported effects are inconsistent among studies^9,10^. In addition, polygenic scores (PGSs) related to depression have been reported to be predictive of side effects that overlap with depressive symptoms, such as suicidality and anxiety^11^. The multi-PGS framework allows combined analysis of multiple relevant PGSs associated with a specific trait or outcome, with the aim of improved prediction accuracy compared to a single PGS^12,13^.

Side effects could be influenced by a combination of many factors spanning attributions, illness representations, personality, clinical, genetic, and social factors. In contrast with traditional statistical models, machine learning methods can optimally be used for prediction analyses involving large datasets that show complex interrelationships and are very often high-dimensional. Machine learning can effectively capture both linear and non-linear relationships across multiple data sources^14,15^. These methods employ data-resampling procedures in holdout samples to estimate models’ parameters and to internally validate inter-variable relationships in unseen individuals. To achieve generalisation, models are validated in independent samples not used for model building. Published machine learning models have used demographics, diagnoses, laboratory values and medication information across various drugs to predict side effects in clinical trials and electronic health record data^16^. For antidepressants, treatment response and improvement in depression severity have been mostly studied within groups of individuals receiving the same treatment^17^. However, few studies have investigated side effect prediction combining genetic and clinical measures^18^.

In this study, we leveraged self-reported questionnaires and genome-wide genetic data from the Genetic Links to Anxiety and Depression Study (GLAD)^19^ to build prediction models of side effect occurrence for three common antidepressant classes: SSRIs, SNRIs, and TCAs. Self-reported side effects and discontinuation (due to side effects) were obtained from the first and last (most recent) year of prescription. We applied six machine learning models incorporating 259 predictors covering five categories: genetic variables, clinical measures, demographic characteristics, illness information, and antidepressant history (Figure 1). We also presented the prevalence, correlation, and association with antidepressant classes in 30 common side effect symptoms. By integrating data on multiple antidepressants in a large cohort, we aimed to provide a more realistic portrayal of real-world drug prescriptions and enhance the potential for early and accurate detection of side effects in personalised antidepressant prescriptions.

**Figure 1.**
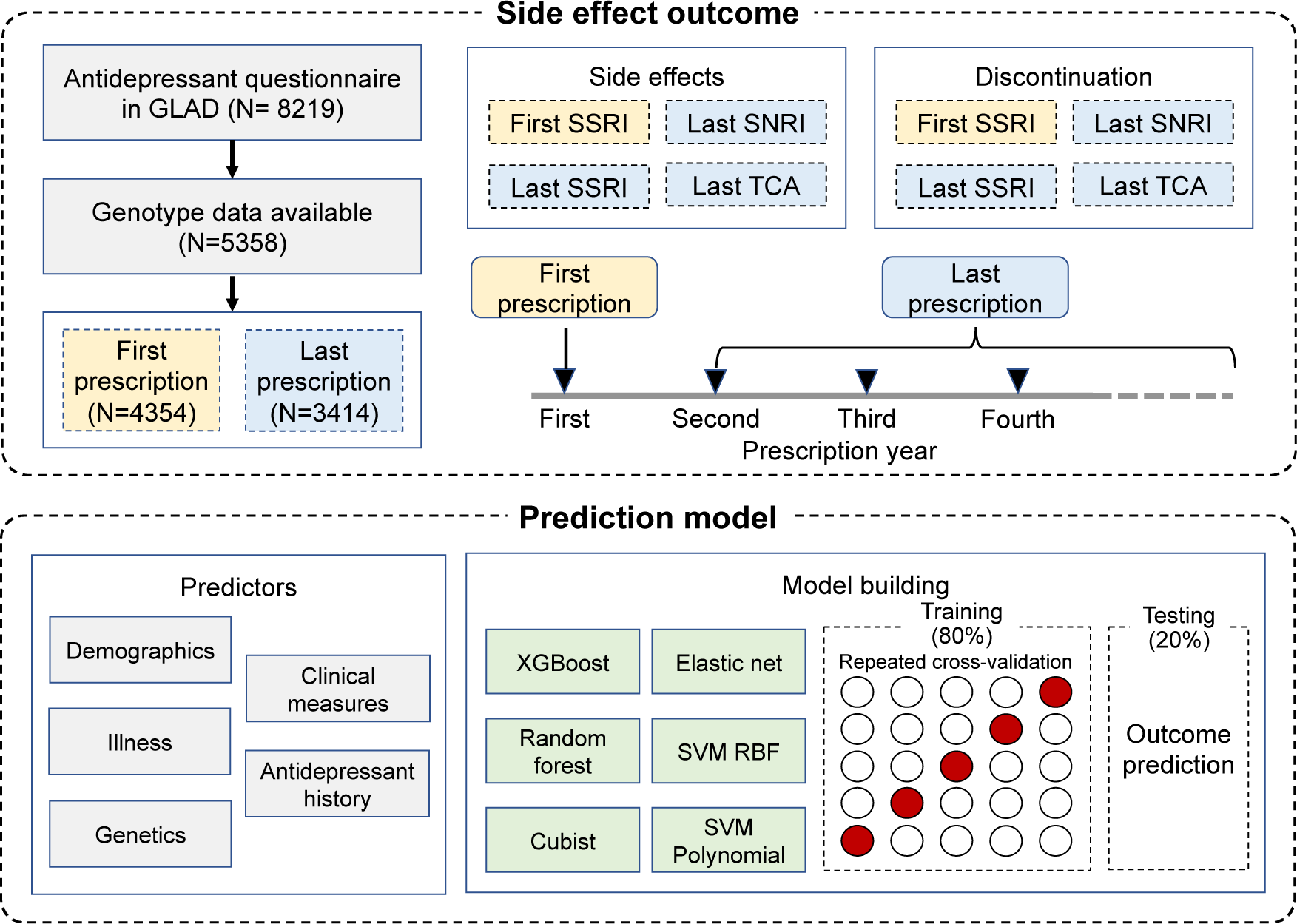
Side effect outcome and prediction models.

## Methods

### Study population

All participants in this study were from the Genetic Links to Anxiety and Depression (GLAD) Study. In summary, individuals from the general population in the UK were recruited using a combination of traditional and social media, as well as clinic-based recruitment, based on the eligibility criterion of meeting a DSM-5 diagnosis for major depressive disorder (MDD) or any anxiety disorder. Participants were invited to complete online sign-up questionnaires, covering demographics, illness, and psychological and behavioural phenotypes relevant to anxiety and depression. Subsequently, participants filled out additional questionnaires that more deeply detailed medication use, psychiatric disorders beyond depression and anxiety, lifestyle, and personal history. A more comprehensive description can be found in Davies et al^19^. GLAD received ethical approval from the London - Fulham Research Ethics Committee (REC reference: 18/LO/1218), while the NIHR BioResource obtained approval from the East of England - Cambridge Central Committee (REC reference: 17/EE/0025).

### Polygenic score (PGS)

Detailed genotyping and imputation steps can be found in Supplementary Methods. Imputed genotypes were aligned with the Hapmap3 reference panel to remove strand-ambiguous SNPs and for PGS calculation. PGSs were computed using MegaPRS^20^ via the GenoPred pipeline^21^. PGSs were reference standardised: MegaPRS was used to first construct PGS in the 1000 Genome European reference panel, following which the mean and standard deviation of PGS output in GLAD samples were scaled to the mean and standard deviation of the reference. GWAS summary statistics from 102 traits were incorporated for both reference PGSs and GLAD PGSs in this study (Supplementary Table 1). PGSs from the GLAD were further rescaled using the reference PGSs to ensure a comparable distribution with the reference.

### Inferred cytochrome metabolic phenotypes

We selected CYP2C19 and CYP2D6 to examine their pharmacogenetic effects, as suggested by the Clinical Pharmacogenetics Implementation Consortium (CPIC) guideline for SSRIs and Tricyclic antidepressants^22,23^. Phasing was conducted using SHAPEIT4 software for the regions of *CYP2C19* and *CYP2D6* obtained from PharmGKB (https://www.pharmgkb.org/)^24^. Star alleles were identified by SNPs matched between the phased data and CPIC definitions (https://cpicpgx.org/; downloaded June 2022)^25^. Allele function and activity values were annotated to include alleles with no, decreased, normal, and increased function (https://cpicpgx.org/)^22^. Metabolic phenotypes of CYP2C19, including poor, intermediate, normal, rapid/ultrarapid, were inferred from genotypes, and classified based on CPIC guidelines. CYP2D6 phenotypes (poor, intermediate, normal) were defined by the consensus recommendations of CPIC and the Dutch Pharmacogenetics Working Group (DPWG)^26^. Since structural variants of *CYP2D6* were unavailable in the imputed genotype data, no rapid/ultrarapid metabolisers were identified. The activity score was determined by summing the activity values of the two star alleles in each individual.

### Outcomes

Participants provided self-reported side effect data for commonly prescribed antidepressants in the UK, including eight selective serotonin reuptake inhibitors (SSRIs), 2 serotonin and norepinephrine reuptake inhibitors (SNRIs), and 10 tricyclic antidepressants (TCAs) (Supplementary Table 2). Participants were asked about any experienced side effects from the prescribed antidepressants and then selected one or more side effects from a list of 30 common symptoms, including *dry mouth, sweating, nausea, vomiting, diarrhoea, constipation, headache, dizziness, memory problems, attention/concentration difficulties, shaking, muscle pain, sleepiness/drowsiness, difficulty getting to sleep, increased anxiety, fast heartbeat, restlessness/agitation, fatigue or weakness, changes in appetite, weight gain, weight loss, itching, rash, runny nose, reduced sexual desire/function, menstrual problems/irregularities, blurred vision, suicidal thoughts, attempted suicide, and others*. Discontinuation was indicated by the response to whether the participant had to stop taking the antidepressant due to side effects.

Antidepressants were ordered by the age at which they were prescribed for each participant. In this study, we included antidepressants prescribed in the first and the most recent (last) year. Information on side effects and discontinuation due to side effects were collected for each antidepressant of each individual. To minimise self-report errors, we limited participants to those who reported at most three different antidepressants during both the first and the last year. Specifically, for first year antidepressants, we focused on whether each participant experienced any side effects or discontinuation with SSRI drugs. For last year antidepressants, we considered the occurrence of side effects and discontinuation for SSRI, SNRI, and TCA antidepressants. In cases where participants were prescribed more than one antidepressant within the same class in either year, side effects and discontinuation were counted if they occurred with either one of the drugs.

### Predictors

We curated 259 items across five categories, including demographics, illness, clinical evaluation, antidepressant history, and genetics, to serve as predictor variables. Core demographic details (e.g., sex, employment status, years of education, marital status, BMI) and health conditions (neurological, allergic, cardiovascular, respiratory, digestive, inflammatory, cancer-related) were obtained from the sign-up questionnaire. Clinical and psychiatric symptoms were assessed through various measures, including lifetime measures of MDD and anxiety (adapted from the Composite International Diagnostic Interview - Short Form (CIDI-SF)), Standardised Assessment of the Severity of Personality Disorders (SASPD), and lifetime assessment of panic disorder (adapted from DSM-5/ICD-11 checklists). In terms of antidepressant histories, we considered antidepressant initiation and last prescription age, the total number and duration of antidepressants taken, and the average number of side effect occurrence, discontinuation, and efficacy (effectiveness of each antidepressant) calculated from all antidepressants taken before the last year. Genetic factors included PGSs for 102 traits and cytochrome metabolic phenotypes, as described above. The complete list of predictors was provided in Supplementary Table 3.

To ensure all predictors in the model have been characterised before the outcome, we chose predictors that preceded the antidepressant initiation including all genetic variables, sex, ethnicity, antidepressant initiation age, lifetime depression and anxiety symptoms to predict outcomes of antidepressants taken in the first year. All predictors were added to the model for the prediction of antidepressants taken in the last year.

### Prediction modelling

#### – Data processing

We removed predictors with missing values > 30%. The remaining were imputed using the random forest algorithm in the MICE package^27^. Predictors with either zero variance or a correlation higher than 0.8 with another predictor were excluded. For categorical factors with more than two categories, they were converted into a set of binary dummy variables in the elastic net and SVM models. Normalisation was applied to each predictor to achieve a standard deviation of one and a mean of zero before training the SVM.

#### -- Model building

We applied six prediction models, including elastic net, support vector machine (SVM) with radial basis function (RBF) and polynomial kernels, XGBoost, random forest, and cubist in this study. A stratified split was employed based on the outcome, randomly allocating 80% for the training data and the remaining 20% for the testing data, maintaining the same outcome proportions as in the original population in each split. Within the training data, a 5- fold stratified cross-validation with 5 repeats were conducted, assigning 20% of the data as a validation set for hyperparameter prioritisation and the remainder for model training. The receiver operating characteristic area under the curve (AUROC) was assessed in the validation set to identify models that maximise performance. The best performing models, along with their hyperparameter combinations, were extracted and employed to predict the outcome class in the test data. Beyond AUROC, we reported accuracy and precision-recall area under the curve (AUPRC) of test data for each model. After model training, a logistic regression was fitted between the probability estimates and the true probabilities to produce the calibrated estimates. More description of each model and data processing can be found in Supplementary Methods.

#### -- Feature importance

For every predictor category (genetics, demographics, illness, clinical and antidepressant information) in the outcomes of drug prescriptions during the last year, we evaluated their importance by excluding the corresponding category of predictors and retrained the model using the same strategy for model building and estimation as described above. The relative reduction in AUROC was calculated for each predictor category compared to the models with the complete set of predictors.

#### -- Prediction across antidepressant classes

For each antidepressant class in the last year of prescription, we compared the model performance between the same drug (training and testing data within the same drug class) and cross drug (training and testing data across different drug classes) analyses to determine the specificity of the model to the drug class. The models with the best performance, identified through the same drug analysis, were then tested on data from other drugs. The Kruskal-Wallis test was used to examine the differences in AUROC values obtained from the same antidepressant class and cross-class analyses.

### Association of side effects with predictors at the last prescription

To further make each predictor’s effect more interpretable and understandable, a univariate logistic regression was performed between each predictor and the six outcomes at the last prescription to evaluate their associations. The false discovery rate was used to adjust the statistical significance for each outcome.

### Correlation of side effect symptoms and drug effect difference

Tetrachoric correlations were applied to assess the correlation of side effect symptoms among SSRIs, SNRIs, and TCAs. Each side effect symptom was coded as a binary variable, with 1 indicating the occurrence of the symptom with any prescribed drugs and 0 showing the absence of the symptom. Hierarchical clustering analysis measured by Euclidean distance was conducted on the distance matrix derived from the correlation matrix of the symptoms. A generalised linear mixed model was constructed to investigate the impact of drug class on each side effect symptom, with drug class as a fixed variable and individual ID as a random variable. SSRIs were set as the reference for comparison with SNRIs and TCAs. Statistical significance was adjusted using the Bonferroni test across the 30 symptoms.

All prediction modelling and association analyses were performed using R v4.2.3^28^.

## Results

### Participant characteristics

All participants in this study were part of the Genetic Links to Anxiety and Depression (GLAD) Study. Overall, 8219 individuals completed the antidepressant questionnaire, and 5358 had available genetic data. In the first year of prescription, most individuals were prescribed SSRIs, with 45% continuing SSRIs and 19.0% transitioning to other antidepressants in the second prescription year (Supplementary Figure 1).

The average prevalence of side effects and discontinuation during the initial five prescription years was 74% and 29%, respectively (Figure 2). In the first year, 6% of participants who initially experienced side effects reported none in the second year. Conversely, 20% of participants without side effects in the first year experienced at least one side effect symptom in the second year (Supplementary Table 4). For the discontinuation outcome, 33% of participants transitioned from discontinuation to no discontinuation in the first two years of prescription. Approximately 8% of participants experienced discontinuation in the second prescription year after having no discontinuation in the first year (Supplementary Table 4).

**Figure 2.**
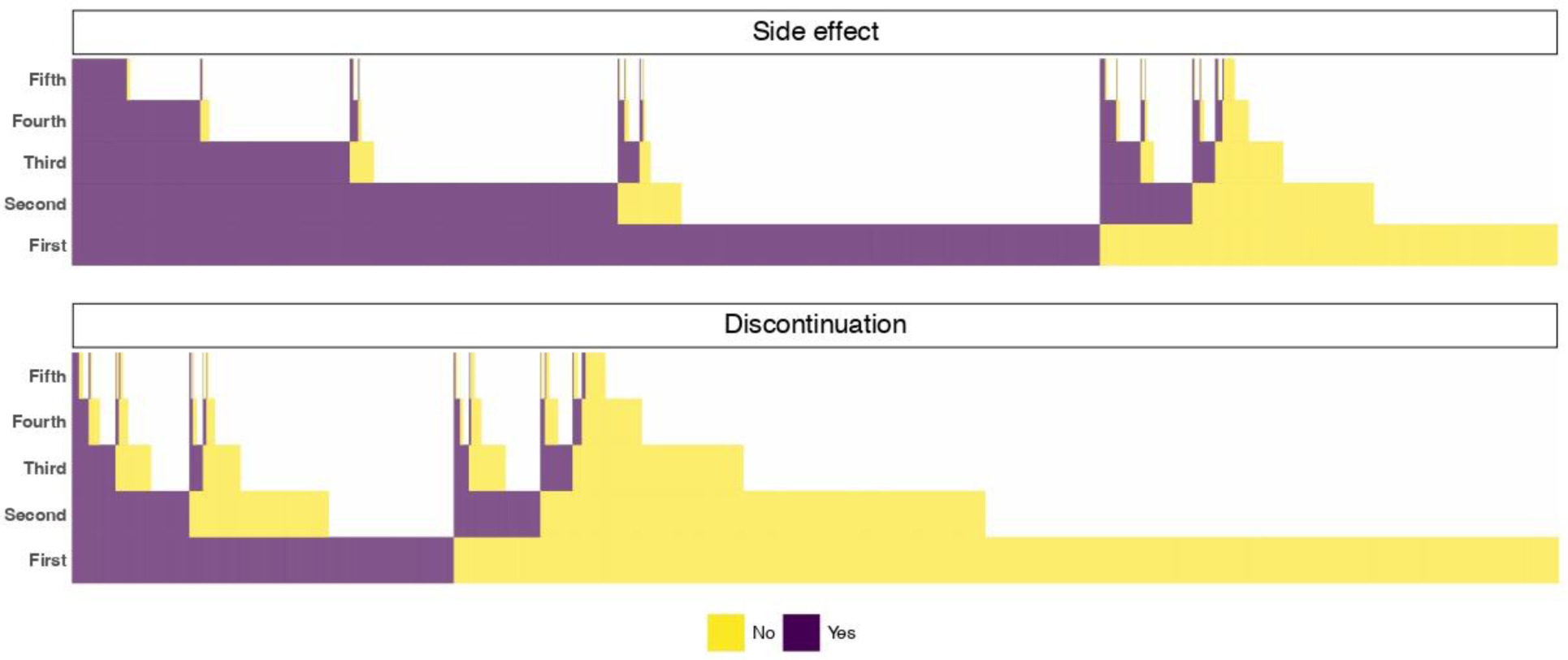
Side effects and discontinuation occurrence at the first five years of prescription, where the x-axis represented each individual (ordered by yes first), and the y-axis showed the time of prescription.

For the prediction models, we included 4354 individuals for SSRIs in the first year of prescription. In addition, 3414 had data for an antidepressant prescription in the last year, comprising 60% (2036) on SSRIs, 22% (753) on SNRIs, and 29% (982) on TCAs. In the initial year, 73% of participants taking SSRIs experienced side effects, and 32% reported discontinuation. In the last year, the reported side effects prevalence for SSRIs was 75%, lower than SNRIs (84%) and TCAs (76%). Discontinuation was highest for TCAs (37%), followed by SSRIs (20.7%) and SNRIs (20.6%) (Table 1).

**Table 1.**
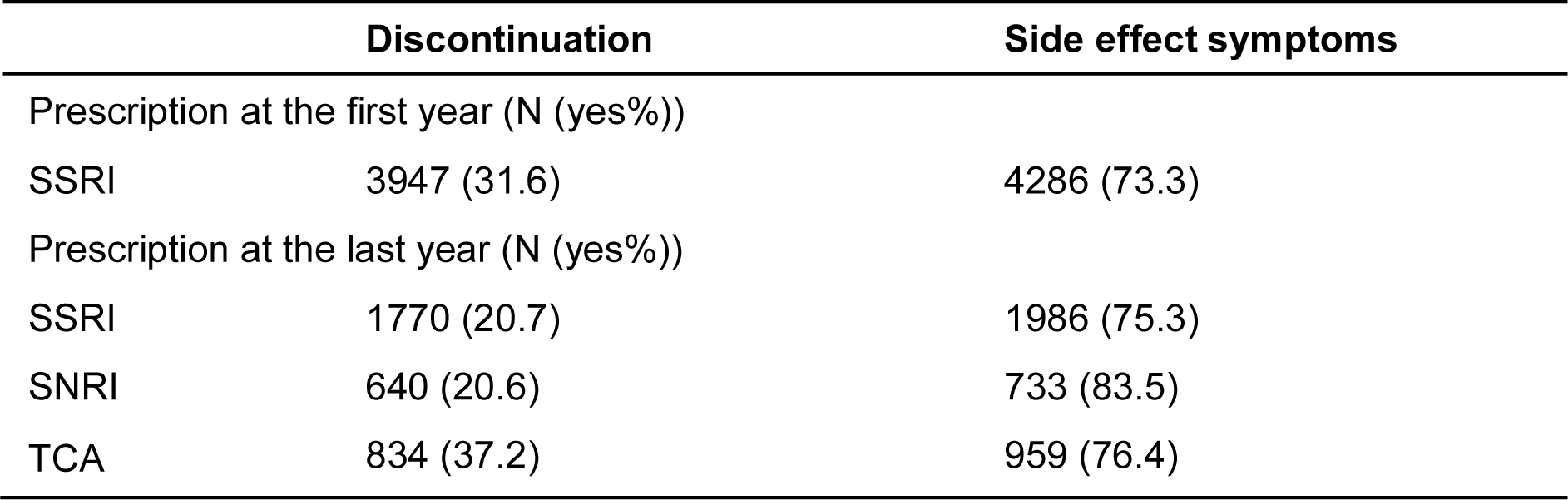
Side effects and discontinuation at the prescription of the first and last year.

### Prediction of antidepressant side effects and discontinuation

In the initial year of SSRIs prescription, the best AUROCs for predicting discontinuation and side effects were 0.65 and 0.60, respectively, achieved by XGBoost (Figure 3a). In the last year of SSRIs prescription, the highest AUROC observed for discontinuation was 0.73, and 0.87 for side effects (Figure 3a). Models including elastic net, random forest and XGBoost achieved AUROC values around 0.70 for discontinuation and above 0.85 for side effects. For SNRIs, the best AUROC was observed in random forest, with 0.71 for discontinuation and 0.80 for side effects. The best models for TCAs had AUROC of 0.71 and 0.87 for predicting discontinuation and side effects, respectively (Supplementary Figure 2). More detailed results for AUROC, AUPRC and accuracy were provided in Supplementary Table 5.

**Figure 3.**
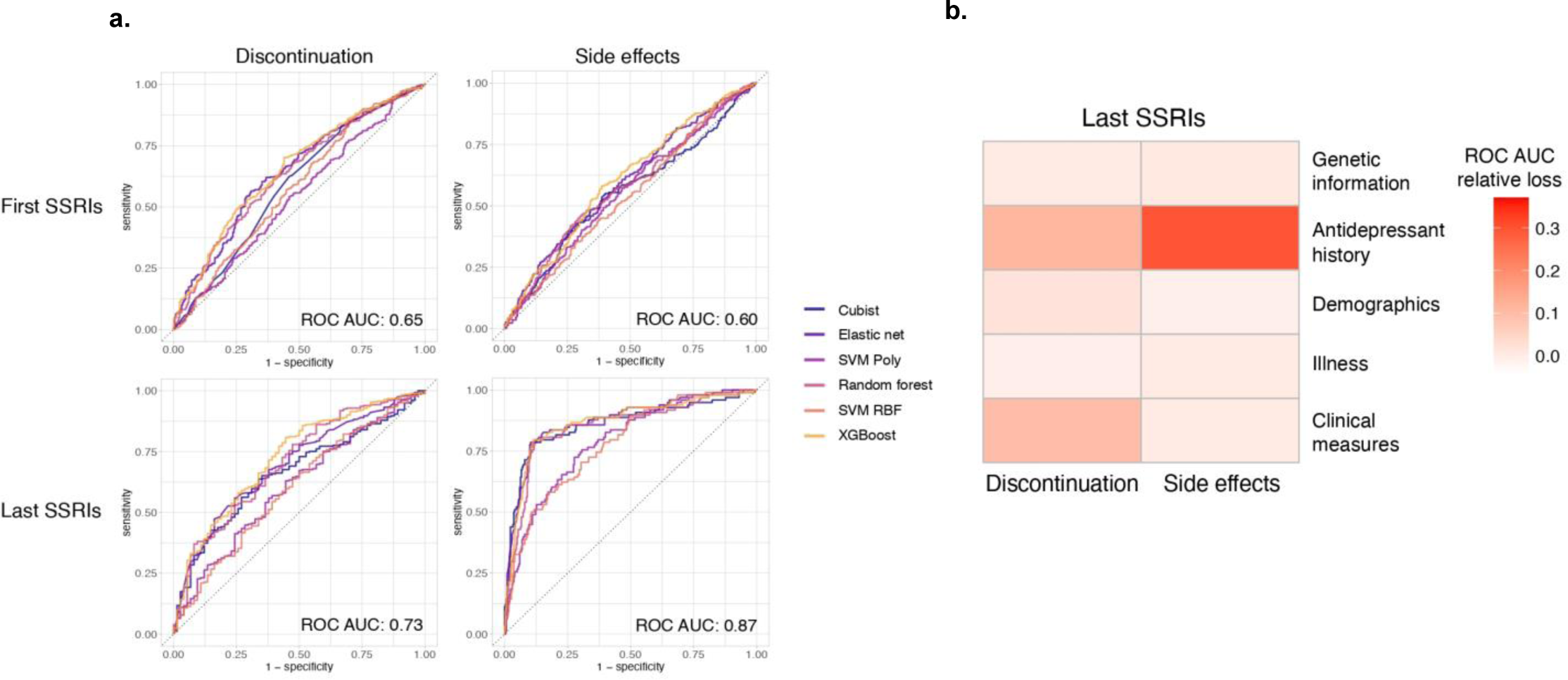
ROC curve (a) and feature importance (b) of SSRIs discontinuation and side effects. Antidepressant history included information of antidepressant initiation and last prescription age, total number and duration of prescribed antidepressants, and the average number of efficacy ratings, side effect occurrence, and discontinuation of all antidepressants taken before the last year of prescription.

For the last year of prescription, we performed cross-class analysis to assess model specificity, comparing the performance of the same training data when tested on different antidepressant classes. For both discontinuation and side effects, the Kruskal-Wallis test revealed no statistically significant differences between the same antidepressant class and different classes for any drug-derived models. This suggested that these models were nonspecific to antidepressant class in the last year of prescription (Supplementary Figure 3).

### Feature importance at the prescription of last year

Next, for each drug class in the last prescription, we evaluated the importance of each predictor category by removing corresponding categories one at a time and retrained the model selected by the best performance of AUROC. We chose XGBoost in this step given it showed superior performance on average across all outcomes described above. Overall, antidepressant history emerged as the most influential category in predicting discontinuation and side effects during the last year of prescription, with a relative reduction in AUROC ranging from 11.4% to 24.0% for discontinuation and 20.4% to 37.1% for side effects (Figure 3b, Supplementary Figure 4). Other clinical measures made a modest contribution to the model for discontinuation, with an impact of 10.0% in SSRIs, 1.4% in SNRIs, and 3.0% in TCAs. Their impact on side effects was considerably less, with the highest relative AUROC reduction of 1.0% in TCA drugs. Genetic factors exhibited the highest impact (1.1%) in predicting side effects and 0.3% for discontinuation (Supplementary Figure 4).

### Association at the prescription of last year

We also conducted univariate logistic regression to examine the association of side effects and discontinuation outcomes with individual features during the last year of prescription. As expected, we observed a strong association between side effect/ discontinuation history and both outcomes across all antidepressant classes (Supplementary Figure 6). For the discontinuation outcome, clinical measures related to depression (low mood, number of episodes, thoughts about death, group/talking therapy), anxiety (anxiety symptoms, duration of worry), and panic disorder (choking, strong fears of attack situations, panic symptoms, medical conditions) were linked to SSRIs, while no significance was observed in other drugs. For side effects, antidepressant prescription age and panic disorders (including panic symptoms, feeling hot or cold) showed associations across all drugs. Other features, including marital status, educational attainment, depressive symptoms, and panic symptoms (strong fears of attack situations, fear of losing control or going crazy, sweating) were associated with any two of the three antidepressant classes (Supplementary Figure 6).

### Side effect symptoms correlation and drug specificity

To investigate the correlations among side effect symptoms and identify how they differed across drug classes, we collected 30 common symptoms from the self-reported questionnaire. In Figure 4, we present the prevalence of side effect symptoms for each drug class across all prescriptions. The most reported side effects included reduced sexual desire, dry mouth, sleepiness/drowsiness, and weight gain, while rash, runny nose, menstrual irregularities, and weight loss had the lowest prevalence. TCAs had most individuals reporting sleepiness/drowsiness, weight gain, dry mouth, and changes in appetite. Reduced sexual desire, nausea, dry mouth, and concentration difficulties were the most common symptoms of SSRIs. Reduced sexual desire, sweating, dry mouth, and nausea were predominantly reported in SNRIs (Supplementary Table 6).

**Figure 4.**
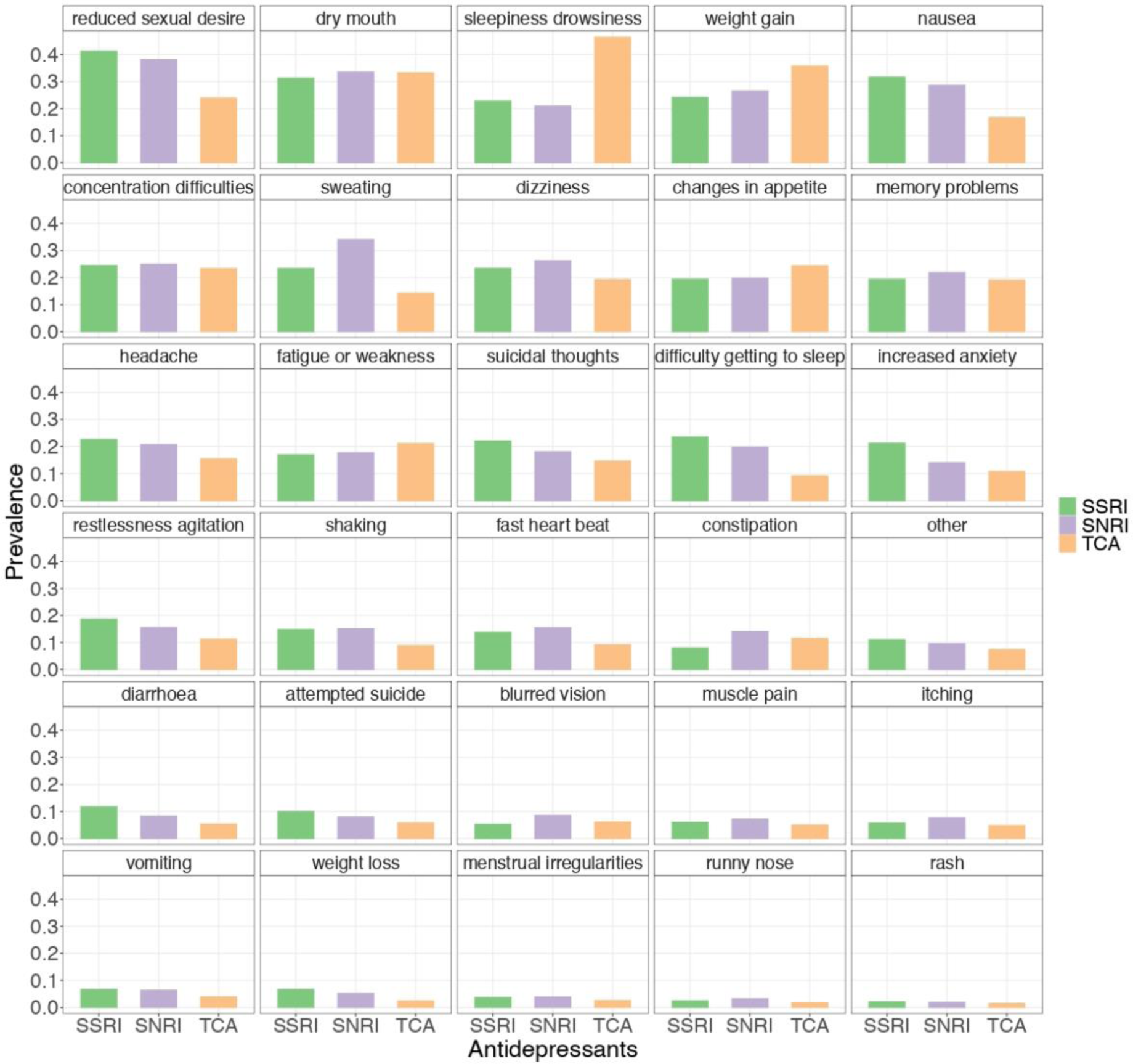
Prevalence of side effect symptoms in antidepressant classes. Symptoms were ordered by their prevalence in the studied population.

For individuals who reported more than one side effect symptom, we examined the correlations between these symptoms across medications. Significant correlations were evident between certain symptoms, such as nausea and vomiting (r = 0.797), memory problems and concentration difficulties (r = 0.803), itching and rash (r = 0.782), as well as suicidal thoughts and attempted suicide (r = 0.861). Moderate correlations were observed between sleepiness/drowsiness and fatigue or weakness (r = 0.663), headache and dizziness (r = 0.653), and a symptom cluster comprising increased anxiety, restlessness/agitation, and fast heart beat (increased anxiety vs restlessness/agitation: r = 0.743; fast heart beat vs restlessness/agitation: r = 0.689; fast heart beat vs increased anxiety: r = 0.639, Supplementary Figure 6).

Finally, we used generalised linear mixed models to investigate the impact of antidepressant classes on each side effect symptom. Compared to SSRIs, SNRIs and TCAs were associated with fewer reports of symptoms like suicidal thoughts, attempted suicide, increased anxiety, diarrhoea, weight loss, vomiting, restlessness/agitation, reduced sexual desire, rash, headache, and difficulty getting to sleep. Conversely, constipation showed a higher occurrence in both SNRIs and TCAs compared to SSRIs. Symptoms such as weight gain, sleepiness/drowsiness, fatigue or weakness, and changes in appetite were more likely to occur in TCAs than SSRIs but were comparable between SNRIs and SSRIs. Dry mouth and concentration difficulties showed no significant differences across all drugs (Figure 5, Supplementary Table 7).

**Figure 5.**
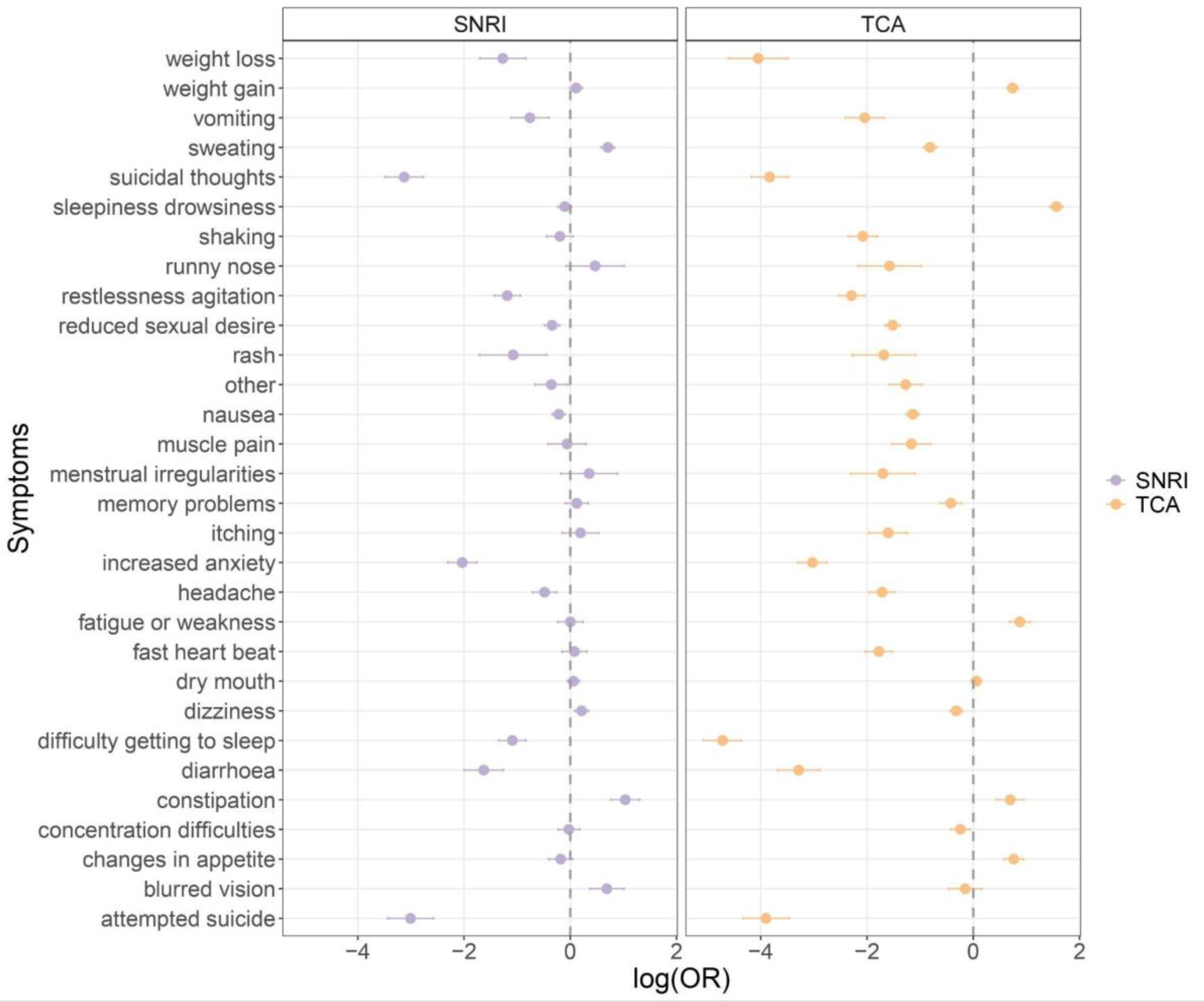
Association of side effect symptoms with drug classes using SSRIs as reference. Each bar represents the effect of SNRIs/TCAs on side effect symptoms compared to SSRIs with 95% confidence intervals.

## Discussion

In this study, we find evidence that self-reported side effects and discontinuation (due to side effects) can be predicted using questionnaires and genetic information, particularly for the most recent antidepressant treatments. Among all predictors, antidepressant history including side effects and discontinuation history showed the primary contribution, indicating individuals tend to have consistent side effect occurrence across different prescriptions. When examining 30 common antidepressant side effect symptoms, the most prevalent symptoms were lower sexual desire, dry mouth, and sleepiness/drowsiness. Most of the 30 symptoms were differentially prevalent between antidepressant classes.

Using machine learning, our results confirmed predicting side effect occurrence was more accurate for the last antidepressant prescription compared to the initial treatment, particularly when considering antidepressant history. Relatively few studies have investigated how drug history influences adverse reactions to the current treatment. However, similar findings are reported in the available literature. For example, the risk of gastrointestinal side effects in nonsteroidal anti-inflammatory drugs (NSAIDs) could be increased if previous NSAID intolerance was identified^29^. In hospitalised older patients, a history of experiencing harmful outcomes from medications in the preceding year was shown to predict adverse drug effects from current medications^30^. A study on treatment-resistant depression also found that a history of electroconvulsive therapy and unsuccessful antidepressant trials were associated with the outcome of vagus nerve stimulation^31^. Despite this, medication history of side effects and discontinuation has been largely overlooked in prediction studies.

Antidepressant side effects have been linked to the severity of depression, polypharmacy, and comorbid status^8,32,33^. Similarly, we observed that clinical measures such as a higher score of depression/panic symptoms and number of episodes could increase the risk of side effects and discontinuation, but their effects were less pronounced than the impact of side effects or discontinuation history. We also identified a large correlation of side effect history among antidepressant classes above 0.62 (SSRIs vs SNRIs: r = 0.66, SSRIs vs TCAs: r = 0.68, SNRIs vs TCAs: r = 0.62), suggesting the occurrence of self-reported side effects might be underlined by other factors regardless of antidepressant class, such as pharmacological, or genetic factors, as indicated in other studies^11^. The role of other factors such as expectation bias should also be considered, especially when using self-reported data^33^.

Genetic predictors, including pharmacokinetic features (*CYP2C19* and *CYP2D6* metabolic phenotypes) along with polygenic risk for multiple disorders and traits only marginally improved model performance in the last prescription. This finding aligned with Poweleit et al.^18^, where the predictive ability of CYP2C19 phenotypes was limited compared to pharmacokinetic parameters including clearance, half-life for escitalopram and sertraline side effects. CYP2C19 poor metabolisers exhibited an OR of 1.99 for the occurrence of the last SSRI side effect in our univariate association analysis, showing a comparable effect size to other studies^10,34^. However, statistical significance was not achieved due to limited power. We did not find an association between CYP2D6 metabolic status and any side effect outcomes and it is important to note that the genotype-determined metabolic phenotype in our study has a lower ability to identify star alleles, particularly for *CYP2D6*. Approximately 7% of *CYP2D6* variants are structural variants, so the star allele calls and metabolic phenotype imputation would be influenced by these missing variants in the genotype leading to the misclassification of metabolic phenotypes and higher false negative findings^25,35^.

We observed a modest predictive ability of PGSs for side effects and discontinuation. Previous studies have reported PGSs for depression, schizophrenia, attention-deficit hyperactivity disorder, and bipolar disorder with treatment response or resistance, but these results mostly yielded no strong evidence of significant associations^36,37^. A study showed PGSs were associated with corresponding side effect symptoms, such as the PGS for depression was linked to depressive symptoms including suicidality and anxiety, and the BMI PGS was associated with weight gain side effects^11^. However, the explained variance attributed to these PGSs (approximately 1%) to predict side effect outcomes was low. Although our study integrated multiple PGS traits to enhance prediction power^12^, we lacked PGSs directly related to antidepressant side effects or responses, which need to be the subject of larger studies^37^. We also acknowledge the need for a larger sample size to fully reflect their predictive ability^38^. Genetic contribution to antidepressant side effects would be further improved with more precise measures of pharmacogenomic phenotypes and sufficient sample size to maximise the PGS evaluation.

As mentioned above, the most commonly reported side effects included reduced sexual desire, dry mouth, sleepiness/drowsiness, and weight gain – consistent with findings from other studies^1,32^. Some symptoms, such as dry mouth, were common in all antidepressant classes, while others, such as reduced sexual desire, were mostly associated with SSRIs or SNRIs, and drowsiness was most prevalent in TCAs. Previous studies have indicated SSRIs were associated with gastrointestinal symptoms, anxiety, insomnia, and sexual side effects, and TCAs broadly conferred an increased risk of weight gain, sedation, sexual dysfunction, and anticholinergic symptoms^4,6,32,39,40^. We confirmed SSRIs were more likely to cause gastrointestinal symptoms (vomiting, diarrhoea, nausea), restlessness agitation, anxiety, weight loss, reduced sexual desire, and difficulty getting to sleep, while individuals taking TCAs experienced more weight gain, sleepiness/drowsiness, change in appetite, and fatigue or weakness. Anticholinergic symptoms such as constipation were more common with TCAs than SSRIs while dry mouth and concentration difficulties showed no differences among drug classes.

We found suicidality (attempted suicide and suicidal thoughts) was mostly reported in SSRIs compared to SNRIs or TCAs. The FDA has added a black box warning for increased suicidality risk among adolescents aged 18-24 with SSRIs prescription^4,41^. However, the effectiveness of this warning is disputed, given increased suicidality among those with severe depression who were not prescribed antidepressants as a result of the warning^42^. While SSRIs were found to increase the risk of suicidality when prescribed for adolescents, the risk was reduced in adults^40^. Among people aged 20 or older when prescribed antidepressants, there was no significant difference in rates of attempted suicide and self-harm between SSRIs and TCAs^43,44^. When stratifying by age, we found that the difference in the rate of attempted suicide between SSRIs and TCAs was more pronounced in individuals younger than 25 than in older participants (Supplementary Table 8). Given the retrospective reporting of prescriptions and side effects in our study of a relatively severe study sample, individuals who had suicidal behaviour after taking antidepressants may have failed to respond leading to a potential ‘survivorship bias’ of our samples. Thus, our results should be interpreted with caution with more evidence needed from other studies.

Our study provides a unique resource to explore the contribution of genetic and medical factors on antidepressant side effects using one of the most powerful and comprehensive self-report records available. Nonetheless, some limitations should be considered. In addition to incomplete metabolic phenotype imputation and limited polygenic prediction due to available PGSs and sample size mentioned above, the retrospective study design with self-report phenotype is prone to subjective interpretations and recall bias. In the GENDEP clinical trial, antidepressant side effects were shown to be reliably measured with self-report instruments, via good agreement with psychiatrists’ ratings. However, this evaluation was limited to the trial period^32^. Linking our data to electronic medical records would allow us to track individuals’ antidepressant trajectories through their lifetime with a more precise assessment compared to self-report^45^. Illness perceptions related to antidepressant medication are not available in our data which may be predictive of treatment adherence or continuation^46^. In addition, our data recorded by age did not distinguish whether antidepressants were prescribed simultaneously or not within the same year. Non-antidepressant medication data was unavailable limiting the evaluation of polypharmacy effects. We also did not collect detailed information on antidepressant dosage, atypical antidepressants (such as Bupropion, Trazodone), or rarer side effects such as myocardial infarction. Finally, our study lacks external validation of models in an independent sample. As our results are primarily based on individuals of European ancestry, further analyses are necessary when generalising our findings to other populations with distinct genetic and social backgrounds.

In conclusion, our study highlighted that self-reported antidepressant side effects and discontinuation can be predicted by incorporating genetic and medical phenotype information, with antidepressant history being a crucial feature. While genetic predictors showed a minor impact, larger samples for discovery GWAS of antidepressant phenotypes and advancements in genetics such as long-read sequencing are needed to fully capture their predictive ability. In our analysis, individuals report experiencing different side effect symptoms across antidepressant classes. Further studies with detailed information on antidepressant regimens and combined treatments will enable a full assessment of antidepressant trajectories and the identification of genetic and clinical risk stratification for potential individualised clinical applications.

## Data Availability

The GLAD study data are available via a data request application to the NIHR BioResource (https://bioresource.nihr.ac.uk/using-our-bioresource/academic-and-clinical-researchers/apply-for-bioresource-data/).

## Funding

This work was supported by the National Institute for Health and Care Research (NIHR) BioResource [RG94028, RG85445], NIHR Biomedical Research Centre [IS-BRC-1215-20018], HSC R&D Division, Public Health Agency [COM/5516/18], MRC Mental Health Data Pathfinder Award (MC_PC_17,217), and the National Centre for Mental Health funding through Health and Care Research Wales. Dr Hübel acknowledges funding from Lundbeckfonden (R276-2018-4581). Johan Källberg Zvrskovec acknowledges funding from the National Institute for Health and Care Research (NIHR) Biomedical Research Centre and Guy’s and St Thomas’ NHS Foundation Trust. Prof McIntosh received funding from the Wellcome Trust (226770/Z/22/Z).

## Conflict of interest

Prof Breen has received honoraria, research or conference grants and consulting fees from Illumina, Otsuka, and COMPASS Pathfinder Ltd. Prof Hotopf is the principal investigator of the RADAR-CNS consortium, an IMI public private partnership, and as such receives research funding from Janssen, UCB, Biogen, Lundbeck and MSD. Prof McIntosh has received research support from Eli Lilly, Janssen, and the Sackler Foundation, and has also received speaker fees from Illumina and Janssen.

## Acknowledgements

We thank the GLAD Study and NIHR BioResource volunteers for their participation and gratefully acknowledge the NIHR BioResource centres, NHS Trusts, and staff for their contribution. We thank the National Institute for Health Research, and Health Data Research UK as part of the Digital Innovation Hub Programme. This study presents independent research funded by the NIHR Biomedical Research Centre at South London Maudsley NHS Foundation Trust and King’s College London. Further information can be found at http://brc.slam.nhs.uk/about/core-facilities/bioresource. The views expressed are those of the authors and not necessarily those of the NHS, the NIHR, the HSC R&D Division, King’s College London, or the Department of Health and Social Care.

## Notes

### Author Declarations

The Genetic Links to Anxiety and Depression (GLAD) Study received ethical approval from the London - Fulham Research Ethics Committee (REC reference: 18/LO/1218), while the NIHR BioResource obtained approval from the East of England - Cambridge Central Committee (REC reference: 17/EE/0025).

