## Supplementary Materials for "Prediction of antidepressant side effects in the Genetic Link to Anxiety and Depression Study"

### Supplementary Figures

Supplementary Figure 1. Antidepressant prescription at the first five years

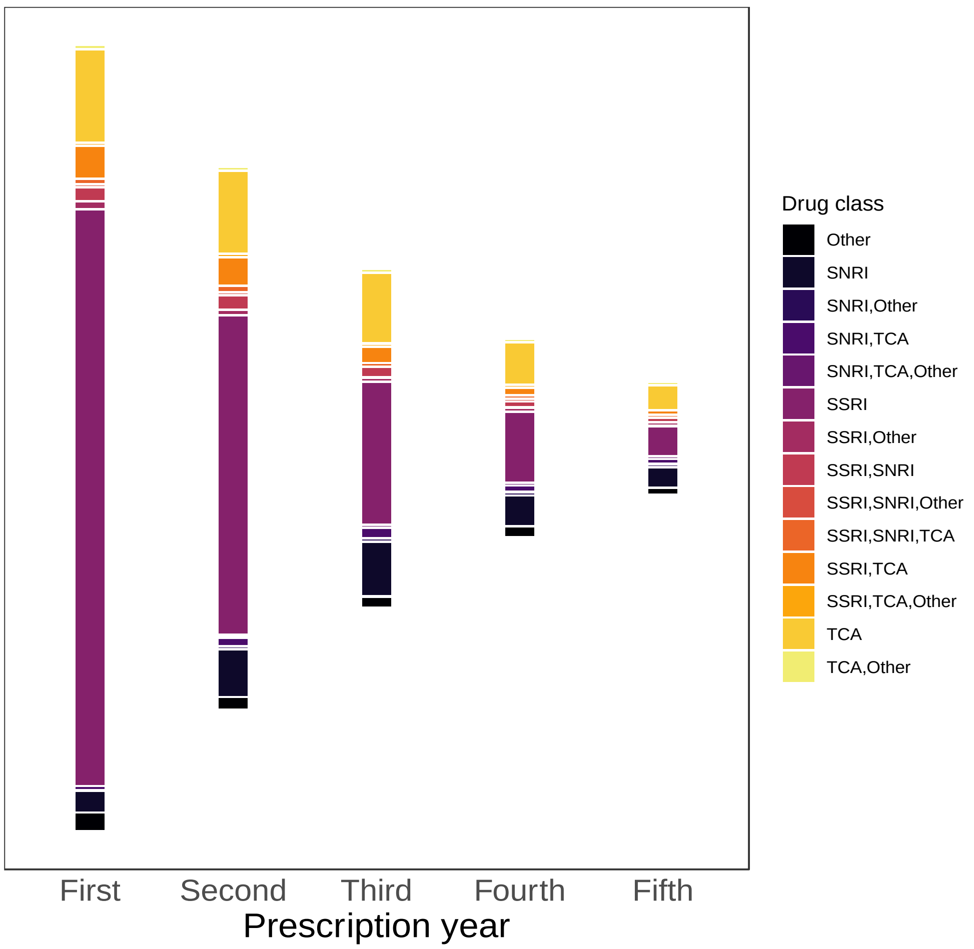

SSRI: selective serotonin reuptake inhibitors; SNRI: serotonin-norepinephrine reuptake inhibitors; TCA: tricyclic antidepressants

Supplementary Figure 2. ROC curve of discontinuation and side effects for SNRIs and TCAs in last year of prescription

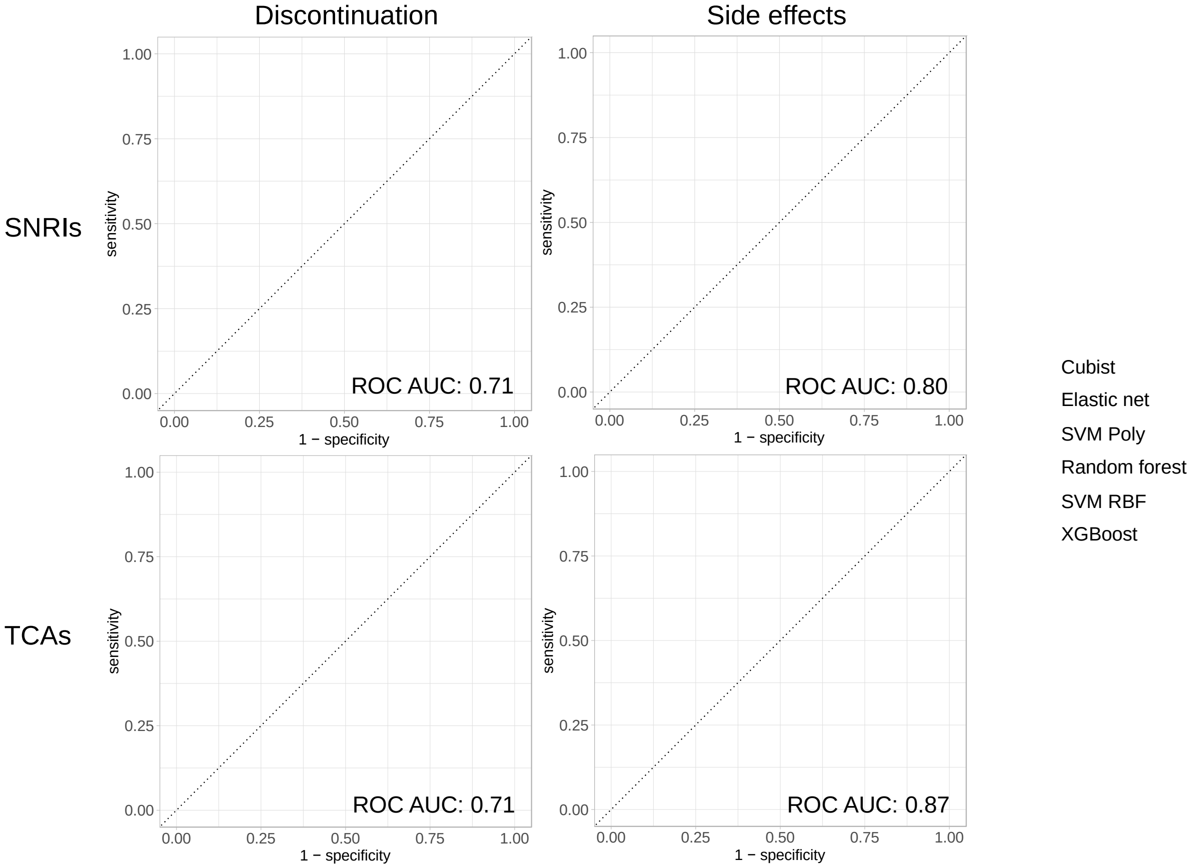

Supplementary Figure 3. Model performance of cross drug analysis

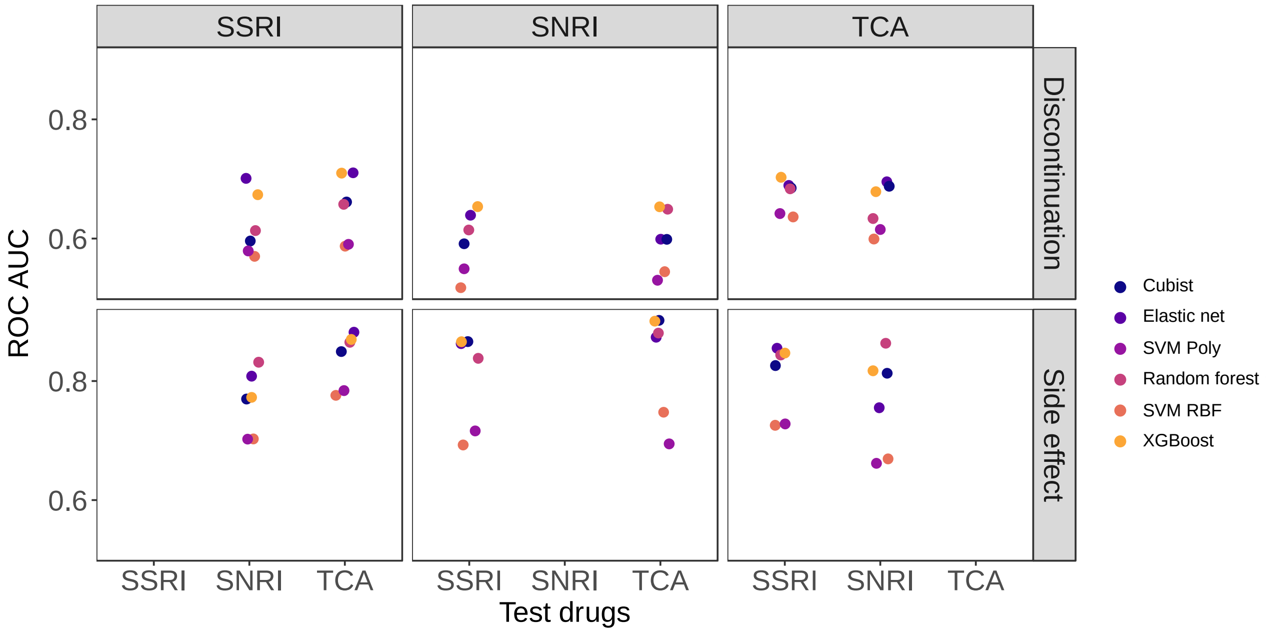

SSRI: selective serotonin reuptake inhibitors; SNRI: serotonin-norepinephrine reuptake inhibitors; TCA: tricyclic antidepressants

Supplementary Figure 4. Feature importance of discontinuation and side effects for SNRIs, and TCAs at the last prescription year

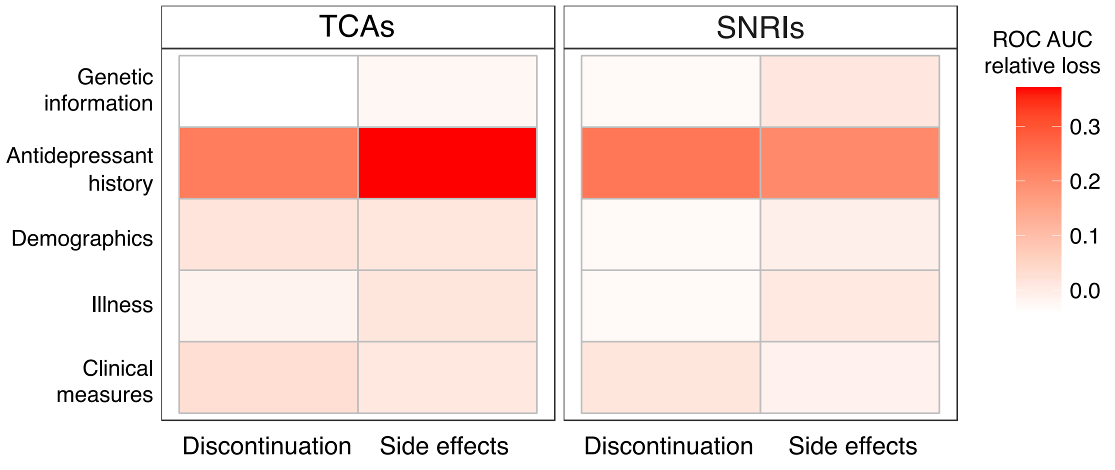

Supplementary Figure 5. Association of discontinuation and side effects with each predictor at the last prescription year

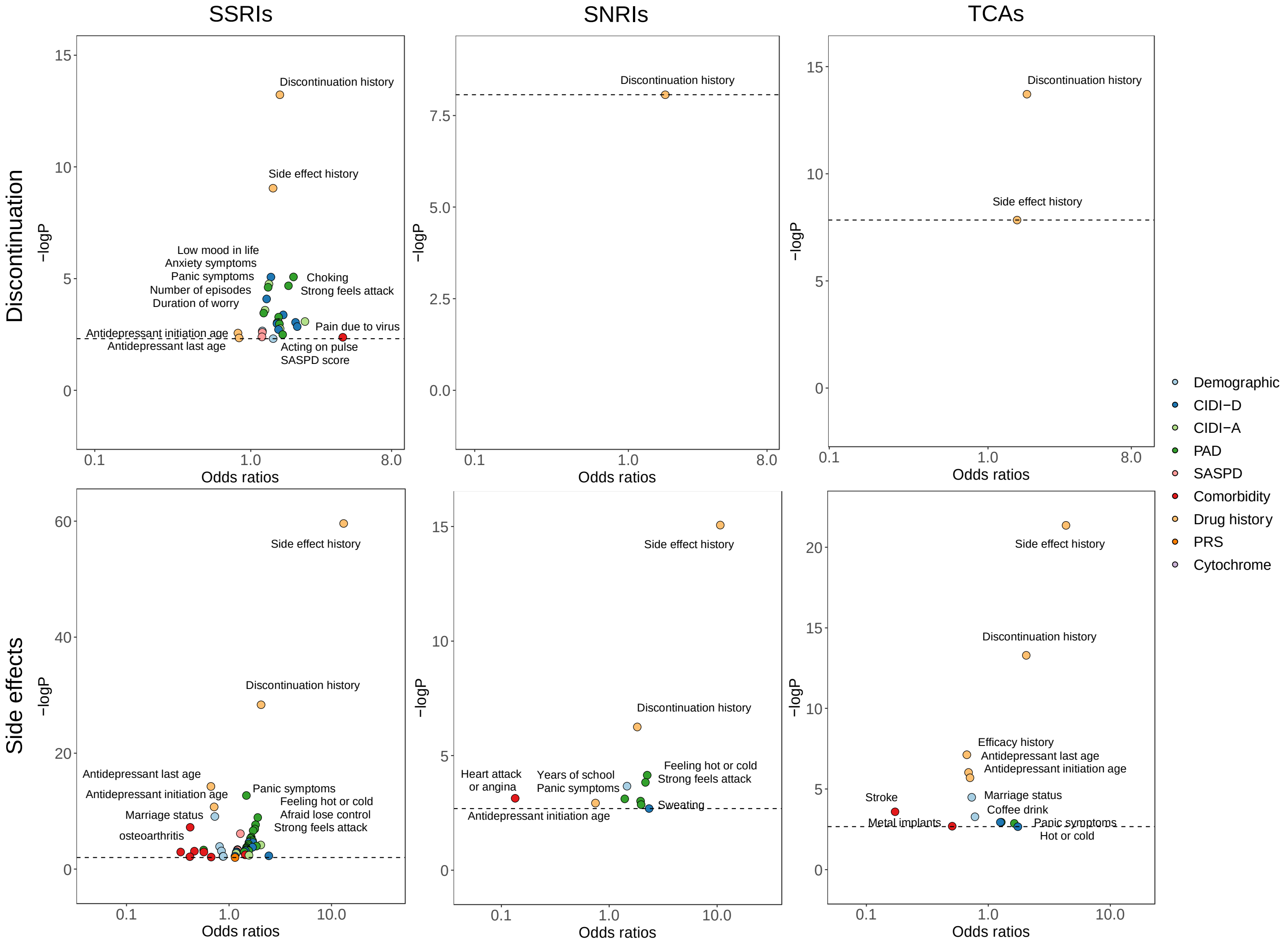

Supplementary Figure 6. Tetrachoric Correlation of side effect symptoms

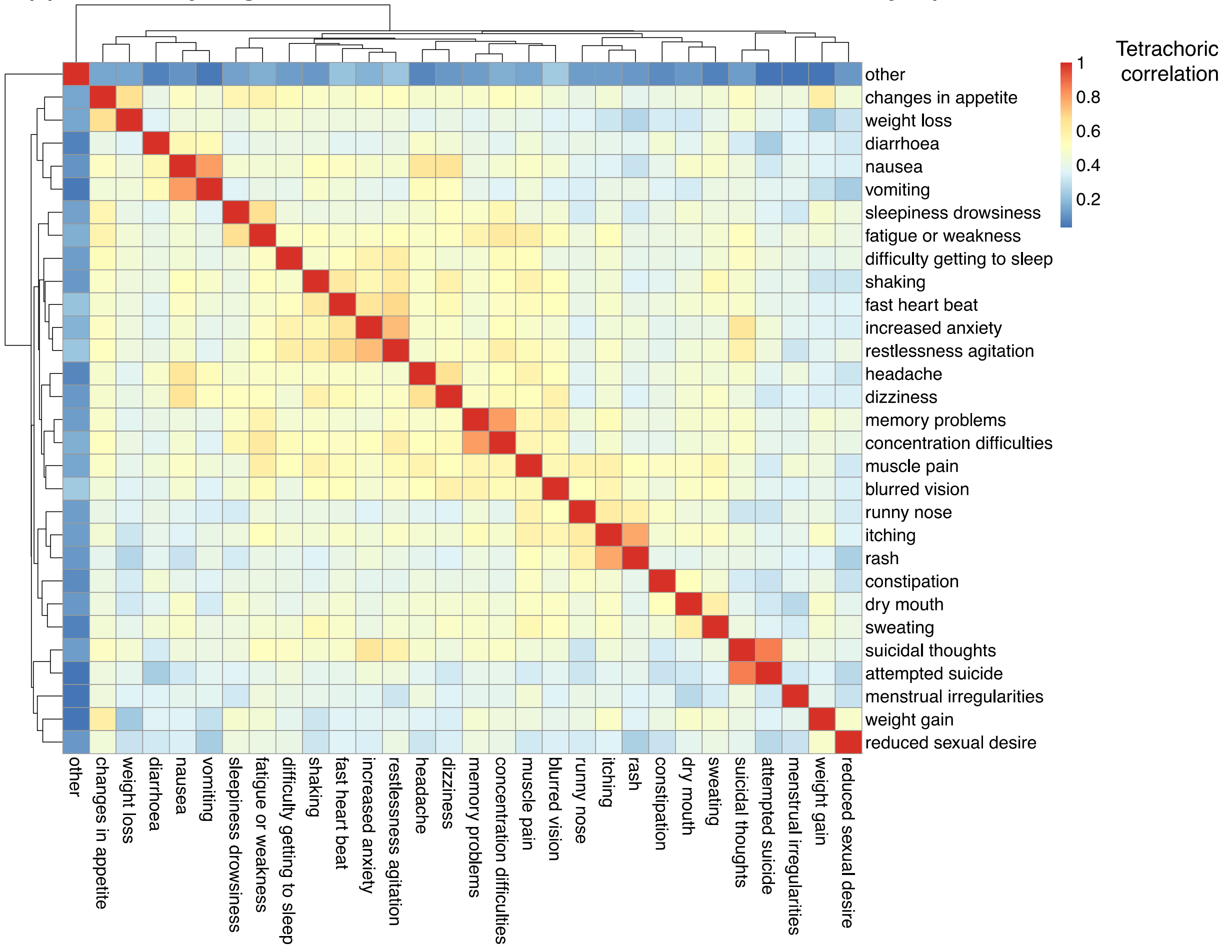

Supplementary Figure 7. Association of side effect symptoms with drug classes using SSRIs as reference.

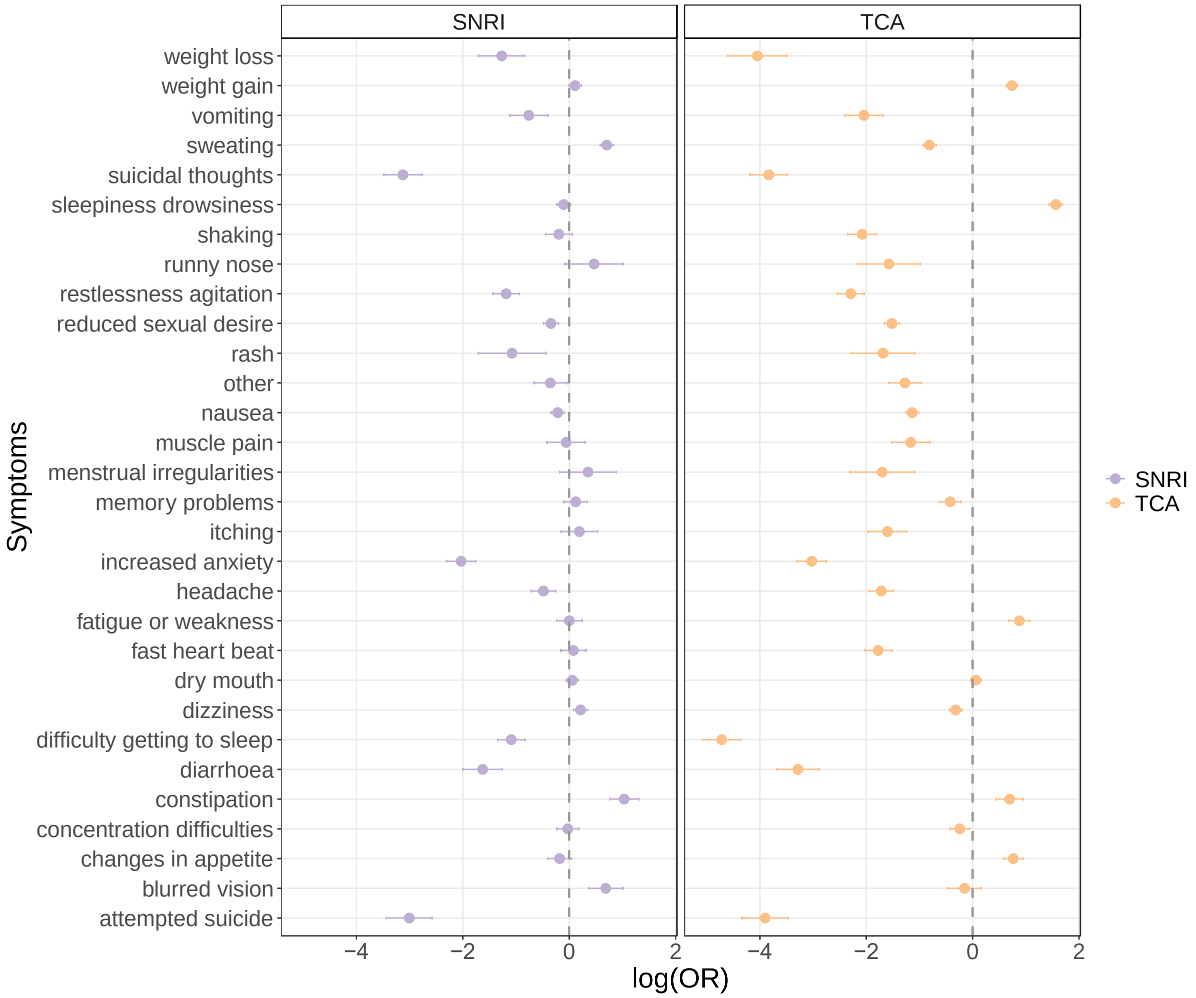

Each bar represents the effect of SNRIs/TCAs on side effect symptoms compared to SSRIs.

### Supplementary Tables

Supplementary Table 2. Antidepressants included in this study

| Antidepressant class | Antidepressants |
|---|---|
| SSRIs | citalopram, fluoxetine, sertraline, paroxetine, escitalopram, fluvoxamine, vortioxetine, dapoxetine |
| SNRIs | venlafaxine, duloxetine |
| TCAs | amitriptyline, dosulepin, clomipramine, nortriptyline, lofepramine, trimipramine, doxepin, imipramine, mirtazapine, mianserin |

SSRIs: selective serotonin reuptake inhibitor
SNRIs: serotonin and norepinephrine reuptake inhibitor
TCAs: tricyclic antidepressant

Supplementary Table 4. Proportion of antidepressant classes in the side effect and discontinuation at the first two years of prescription

| | SSRIs | SNRIs | TCAs |
|---|---|---|---|
| Side effects (first + second) (%) | | | |
| Yes + no | 80.6 | 2.5 | 15.5 |
| No + yes | 60.3 | 2.0 | 22.8 |
| Yes + yes | 80.3 | 3.0 | 15.6 |
| No + no | 75.3 | 2.8 | 17.0 |
| Discontinuation (first + second) (%) | | | |
| Yes + no | 80.1 | 3.0 | 15.6 |
| No + yes | 75.1 | 2.8 | 17.6 |
| Yes + yes | 82.1 | 2.8 | 14.2 |
| No + no | 77.4 | 2.8 | 15.9 |

Supplementary Table 5. Model performance of discontinuation and side effects

| Discontinuation | | | | Side effect | | | |
|---|---|---|---|---|---|---|---|
| | ROC AUC | PRC AUC | Accuracy | | ROC AUC | PRC AUC | Accuracy |
| First SSRIs | | | | First SSRIs | | | |
| Elastic net | 0.64 | 0.60 | 0.69 | Elastic net | 0.59 | 0.78 | 0.74 |
| SVM RBF | 0.58 | 0.63 | 0.69 | SVM RBF | 0.54 | 0.76 | 0.73 |
| SVM poly | 0.54 | 0.64 | 0.69 | SVM poly | 0.55 | 0.76 | 0.73 |
| Cubist | 0.59 | 0.66 | 0.68 | Cubist | 0.55 | 0.73 | 0.73 |
| Random forest | 0.63 | 0.60 | 0.69 | Random forest | 0.58 | 0.77 | 0.73 |
| XGBoost | 0.65 | 0.59 | 0.69 | XGBoost | 0.60 | 0.78 | 0.73 |
| Last SSRIs | | | | Last SSRIs | | | |
| Elastic net | 0.70 | 0.70 | 0.79 | Elastic net | 0.87 | 0.86 | 0.87 |
| SVM RBF | 0.62 | 0.72 | 0.80 | SVM RBF | 0.78 | 0.85 | 0.80 |
| SVM poly | 0.68 | 0.70 | 0.79 | SVM poly | 0.78 | 0.85 | 0.80 |
| Cubist | 0.66 | 0.73 | 0.80 | Cubist | 0.86 | 0.85 | 0.87 |
| Random forest | 0.70 | 0.69 | 0.79 | Random forest | 0.85 | 0.85 | 0.86 |
| XGBoost | 0.73 | 0.68 | 0.80 | XGBoost | 0.87 | 0.86 | 0.86 |
| Last SNRIs | | | | Last SNRIs | | | |
| Elastic net | 0.63 | 0.73 | 0.79 | Elastic net | 0.79 | 0.90 | 0.87 |
| SVM RBF | 0.51 | 0.75 | 0.79 | SVM RBF | 0.69 | 0.88 | 0.84 |
| SVM poly | 0.66 | 0.71 | 0.80 | SVM poly | 0.63 | 0.87 | 0.84 |
| Cubist | 0.63 | 0.74 | 0.79 | Cubist | 0.76 | 0.89 | 0.87 |
| Random forest | 0.71 | 0.69 | 0.80 | Random forest | 0.80 | 0.90 | 0.87 |
| XGBoost | 0.69 | 0.71 | 0.80 | XGBoost | 0.80 | 0.89 | 0.88 |
| Last TCAs | | | | Last TCAs | | | |
| Elastic net | 0.71 | 0.51 | 0.71 | Elastic net | 0.87 | 0.87 | 0.87 |
| SVM RBF | 0.65 | 0.52 | 0.68 | SVM RBF | 0.69 | 0.83 | 0.76 |
| SVM poly | 0.66 | 0.52 | 0.69 | SVM poly | 0.72 | 0.84 | 0.78 |
| Cubist | 0.69 | 0.51 | 0.69 | Cubist | 0.80 | 0.85 | 0.87 |
| Random forest | 0.65 | 0.53 | 0.66 | Random forest | 0.82 | 0.86 | 0.86 |
| XGBoost | 0.70 | 0.50 | 0.69 | XGBoost | 0.84 | 0.86 | 0.87 |

ROC AUC: area under the receiver operating characteristics curve
PRC AUC: area under the precision-recall curve
SVM: support vector machine
RBF: radial basis function
Poly: polynomial

Supplementary Table 6. Prevalence of side effect symptoms in antidepressant classes

| Symptoms | SSRIs | SNRIs | TCAs |
|---|---|---|---|
| Dry mouth | 0.313 | 0.335 | 0.332 |
| Sweating | 0.234 | 0.341 | 0.142 |
| Nausea | 0.317 | 0.287 | 0.167 |
| Vomiting | 0.066 | 0.063 | 0.039 |
| Diarrhoea | 0.117 | 0.083 | 0.053 |
| Constipation | 0.080 | 0.140 | 0.115 |
| Headache | 0.226 | 0.208 | 0.155 |
| Dizziness | 0.234 | 0.262 | 0.193 |
| Memory problems | 0.194 | 0.219 | 0.191 |
| Concentration difficulties | 0.245 | 0.250 | 0.234 |
| Shaking | 0.149 | 0.151 | 0.089 |
| Muscle pain | 0.060 | 0.072 | 0.050 |
| Sleepiness/drowsiness | 0.228 | 0.210 | 0.464 |
| Difficulty getting to sleep | 0.236 | 0.198 | 0.091 |
| Increased anxiety | 0.213 | 0.140 | 0.108 |
| Fast heart beat | 0.138 | 0.155 | 0.092 |
| Restlessness agitation | 0.187 | 0.156 | 0.113 |
| Fatigue or weakness | 0.170 | 0.177 | 0.212 |
| Changes in appetite | 0.194 | 0.198 | 0.244 |
| Weight gain | 0.242 | 0.265 | 0.358 |
| Weight loss | 0.066 | 0.052 | 0.024 |
| Itching | 0.057 | 0.077 | 0.048 |
| Rash | 0.021 | 0.019 | 0.015 |
| Runny nose | 0.024 | 0.032 | 0.018 |
| Reduced sexual desire | 0.412 | 0.382 | 0.240 |
| Menstrual irregularities | 0.037 | 0.038 | 0.025 |
| Blurred vision | 0.052 | 0.085 | 0.061 |
| Suicidal thoughts | 0.221 | 0.181 | 0.146 |
| Attempted suicide | 0.100 | 0.080 | 0.058 |
| Other | 0.111 | 0.096 | 0.075 |

Supplementary Table 7. Association between side effect symptoms and drug class using SSRI as the reference

| Symptoms | Odds ratio | Low 95% CI | High 95% CI | Adjusted P value | Odds ratio | Low 95% CI | High 95% CI | Adjusted P value |
|---|---|---|---|---|---|---|---|---|
| | SNRIs | | | | TCAs | | | |
| Attempted suicide | 0.049 | 0.032 | 0.076 | 3.723E-41 | 0.020 | 0.013 | 0.031 | 5.391E-68 |
| Blurred vision | 1.988 | 1.438 | 2.750 | 9.785E-04 | 0.860 | 0.628 | 1.178 | 1 |
| Concentration difficulties | 0.974 | 0.792 | 1.197 | 1 | 0.785 | 0.654 | 0.941 | 2.654E-01 |
| Changes in appetite | 0.834 | 0.665 | 1.045 | 1 | 2.145 | 1.786 | 2.576 | 1.000E-14 |
| Constipation | 2.814 | 2.141 | 3.698 | 3.477E-12 | 2.005 | 1.555 | 2.585 | 2.462E-06 |
| Diarrhoea | 0.196 | 0.136 | 0.284 | 2.145E-16 | 0.037 | 0.025 | 0.056 | 2.781E-57 |
| Difficulty getting to sleep | 0.336 | 0.260 | 0.434 | 2.017E-15 | 0.009 | 0.006 | 0.013 | 1.329E-145 |
| Dizziness | 1.238 | 1.085 | 1.414 | 4.748E-02 | 0.727 | 0.645 | 0.819 | 5.042E-06 |
| Dry mouth | 1.060 | 0.945 | 1.190 | 1 | 1.061 | 0.962 | 1.171 | 1 |
| Fast heart beat | 1.082 | 0.856 | 1.369 | 1 | 0.169 | 0.131 | 0.219 | 2.711E-40 |
| Fatigue or weakness | 1.001 | 0.787 | 1.273 | 1 | 2.405 | 1.976 | 2.927 | 6.091E-17 |
| Headache | 0.616 | 0.489 | 0.776 | 1.193E-03 | 0.180 | 0.142 | 0.227 | 1.561E-45 |
| Increased anxiety | 0.131 | 0.100 | 0.172 | 1.239E-46 | 0.049 | 0.037 | 0.064 | 1.918E-104 |
| Itching | 1.211 | 0.860 | 1.706 | 1 | 0.201 | 0.141 | 0.289 | 7.594E-17 |
| Memory problems | 1.129 | 0.906 | 1.408 | 1 | 0.656 | 0.536 | 0.804 | 1.426E-03 |
| Menstrual irregularities | 1.427 | 0.836 | 2.434 | 1 | 0.183 | 0.100 | 0.334 | 9.904E-07 |
| Muscle pain | 0.944 | 0.659 | 1.352 | 1 | 0.312 | 0.218 | 0.447 | 6.287E-09 |
| Nausea | 0.805 | 0.710 | 0.911 | 1.815E-02 | 0.321 | 0.283 | 0.363 | 6.250E-71 |
| Other | 0.701 | 0.514 | 0.955 | 7.286E-01 | 0.280 | 0.206 | 0.381 | 1.372E-14 |
| Rash | 0.341 | 0.181 | 0.644 | 2.746E-02 | 0.186 | 0.102 | 0.337 | 9.508E-07 |
| Reduced sexual desire | 0.710 | 0.616 | 0.818 | 6.917E-05 | 0.219 | 0.190 | 0.253 | 2.425E-94 |
| Restlessness agitation | 0.306 | 0.238 | 0.392 | 3.846E-19 | 0.101 | 0.079 | 0.130 | 2.467E-70 |
| Runny nose | 1.597 | 0.924 | 2.760 | 1 | 0.207 | 0.115 | 0.374 | 5.459E-06 |
| Shaking | 0.823 | 0.641 | 1.056 | 1 | 0.125 | 0.095 | 0.164 | 6.685E-49 |
| Sleepiness drowsiness | 0.902 | 0.792 | 1.028 | 1 | 4.776 | 4.220 | 5.405 | 7.002E-134 |
| Suicidal thoughts | 0.044 | 0.031 | 0.063 | 3.410E-63 | 0.022 | 0.015 | 0.031 | 3.147E-100 |
| Sweating | 2.028 | 1.789 | 2.300 | 8.252E-27 | 0.444 | 0.391 | 0.504 | 1.086E-34 |
| Vomiting | 0.468 | 0.328 | 0.669 | 9.281E-04 | 0.129 | 0.090 | 0.186 | 6.655E-27 |
| Weight gain | 1.116 | 0.987 | 1.262 | 1 | 2.095 | 1.887 | 2.326 | 2.431E-42 |
| Weight loss | 0.281 | 0.183 | 0.433 | 2.501E-07 | 0.017 | 0.010 | 0.030 | 9.414E-45 |

Supplementary Table 8. Association between attempted suicide and drug class at the first and the last prescription stratified by age using SSRIs as the reference

| | Odds ratio | Low 95% CI | High 95% CI | p-value |
|---|---|---|---|---|
| First TCAs | | | | |
| < 25 | 0.0011 | 0.00016 | 0.0068 | 6.26E-13 |
| > 25 | 0.058 | 0.011 | 0.302 | 0.000721 |
| Last TCAs | | | | |
| < 25 | 1.7E-06 | 4.85E-08 | 6.13E-05 | 3.23E-13 |
| > 25 | 0.080 | 0.010 | 0.644 | 0.0176 |

### Supplementary Methods

#### Genotyping and imputation

Saliva samples were obtained via the Isohelix saliva DNA sample kit through postal services. Genotyping was conducted using the UK Biobank v2 Axiom array, encompassing over 850,000 genetic variants. Affymetrix software and the UK Biobank pipeline software were used for genotype assignment, and standard quality control procedures were implemented to eliminate unqualified individuals based on criteria such as individual missingness > 5%, and identity by descent (IBD) outliers (3SD from the average). Variants were excluded if they had missingness > 5%, a minor allele frequency (MAF) < 1%, and Hardy-Weinberg equilibrium p-values < $10^{-10}$. The analyses presented were carried in the European ancestry subset as other ancestries were not sufficiently represented. After quality control, imputation was carried out using the TOPMed reference panel (version r2 on GRCh38), and variants were filtered with quality (R2) > 0.3 and MAF > 0.1%.

#### Prediction models

##### -- Elastic net

The elastic net is a linear regularization model that combines the penalties of Ridge and Lasso methods. The Ridge penalty limits the size of the coefficient vector and shrinks correlated coefficients towards one another, while the Lasso penalty introduces sparsity among the coefficients and selects one over the other correlated features. In elastic net, we implemented a grid search strategy to explore every combination of penalty and mixture hyperparameters. The penalty value determined the amount of regularization applied to coefficients to reduce the complexity and overfitting of the prediction model, with 20 candidates ranging equally from 0.0001 to 1. The mixture value represented the tradeoff between Ridge and Lasso penalties, with candidates between 0.1 and 1 in steps of 0.1. To address the class imbalance in the outcome, case weights were introduced to specify the influence of each observation during model estimation. The weights were determined based on the inverse proportion of the outcome class to which each observation belongs.

##### -- SVM

Support Vector Machines (SVM) are models that construct a hyperplane or a set of hyperplanes in high-dimensional space to classify samples into categories. The optimal hyperplane represents the largest margin between the nearest data points, known as support vectors, within two classes. A kernel function is applied to transform the original space into a much higher-dimensional space for effective separation. The hyperparameter grid search for SVM included the cost regularization value, as well as sigma for the radial basis function (RBF) kernel and the degree for the polynomial kernel. The cost parameter regulates the smoothness of the fitted function, including RBF and polynomial kernels, with higher values leading to less smooth functions. We tuned 20 values spanning from 0 to 32 for the cost parameter in the RBF kernel and 50 values in the polynomial kernel. In RBF SVM, sigma represents the inverse kernel width, and we explored 10 values spanning from 1e-10 to 1. In polynomial SVM, the degree parameter specifies the degree of the polynomial function, with integer values from 1 to 3. All combinations of hyperparameter candidates were trained during the model estimation. To address the class imbalance issue, we used the Synthetic Minority Over-sampling Technique upsampling algorithm. This algorithm generates samples from the minority class using nearest neighbours to balance the class proportions.

**--Tree models**

**-- Random forest**

Random forest is an enhancement of the decision tree method that leverages bagging to reduce high variance of individual tree predictions. Through bootstrapping, different subsets are resampled, and the results of each subset are averaged to form an ensemble evaluation. To further decrease correlation among trees within each subset, variables are randomly selected for prediction in each tree. In the random forest model, a design of Latin hypercube grid search was used which is a space-filling parameter approach involving random sampling of points to comprehensively cover the parameter space, ensuring any portion of the space has an observed combination. The hyperparameters grid size was set to 200, with the number of features randomly sampled at each split (ranging from number of features*0.05, to number of features*0.5), the minimal node size (ranging from 2 to 40), and the number of trees (ranging from 1 to 2000).

**-- XGBoost**

XGBoost is an ensemble method designed to enhance the performance of individual trees in parallel and sequentially combine their outputs. It is trained to minimize the residual errors of its predecessors through a gradient descent algorithm, providing a scalable and highly accurate implementation for optimal computing power and model performance. In our application of XGBoost, we utilized a Latin hypercube grid search design with a grid size of 200. This encompassed various hyperparameters, including the number of features randomly sampled at each split (ranging from 0 to the number of features), minimal node size (ranging from 1 to 40), number of trees (trees, ranging from 100 to 1000), depth of trees (ranging from 1 to 15), minimum loss reduction (ranging from -10 to 1.5 on a log-10 scale), learning rate (ranging from -10 to -1 on a log-10 scale), proportion of observations sampled (ranging from 0 to 1), amount of L1 regularization (ranging from -10 to 1 on a log-10 scale), amount of L2 regularization (ranging from -10 to 1 on a log-10 scale), balance of events and non-events (ranging from 0.8 to 1.2), and iterations before stopping (ranging from 3 to 20).

**-- Cubist**

Cubist models represent a form of decision tree modelling that incorporates rules to segment the data. This algorithm involves two key steps: the first step establishes a set of rules to divide the data into smaller subsets, and the second step applies a regression model to these subsets to make predictions. In our application of cubist, we employed a Latin hypercube grid search design with a grid size of 200. This design combined parameters involving combinations of tree numbers (ranging from 1 to 2000) and minimum node size (ranging from 2 to 40).

Similar to the elastic net, case weights were assigned in each tree model to signify the influence of each observation during model estimation, determined by the inverse proportion of the outcome class.
